# International Consensus on Sports, Exercise, and Physical Activity participation during post-operative interventions for Adolescent Idiopathic Scoliosis: an e-Delphi study

**DOI:** 10.1101/2025.03.23.25324479

**Authors:** Susanna Tucker, Nicola R Heneghan, Alison Rushton, Adrian Gardner, Emily Russell, Andrew Soundy

## Abstract

**Introduction:** Physiotherapists and surgeons have a significant role in promoting participation and offering a graded return to sports, exercise, and physical activity following spinal fusion in adolescent idiopathic scoliosis (AIS). However, there is a lack of evidence to guide post-operative rehabilitation and variability worldwide. This study aims to obtain consensus on 1) when it is safe and 2) how an individual with AIS might return to sports, exercise, and physical activity.

**Methods and analysis:** An international electronic 3 round Delphi study was conducted and reported. Eligible expert surgeons or physiotherapists had either specific clinical or research experience in AIS. Round 1 included a series of open-ended questions. Round 2 provided a summary of the existing literature for participants to review and rate items. Round 3 participants were asked to re-rate responses. Consensus was determined through content analysis of open statements >1 participant, round 2 statements achieving >75% on a 5-point Likert scale, and round 3 Kendall’s coefficient of concordance.

**Results:** From 53 recruited participants (18 countries, 1 unknown), 41 responded to round 1, 32 to round 2, and 29 to Round 3 (14 surgeons, 15 physiotherapists). Round 1 generated 85 statements under 19 themes surrounding graded return to sports, rehabilitation milestones, philosophical approaches, and treatment modalities. Round 2 generated 63 statements with >75% agreement. Round 2 open comments generated 22 statements. The 66 statements agreed in round 3 generated 9 themes with corresponding statements regarding different phases of care. All round 3 statements demonstrated significant (p<0.001) moderate agreement (W=0.5). A Wilcoxon Sum-rank result (p<0.05) showed stability between rounds 2 and 3. An additional 5 statements (total 71 statements) were generated from round 3 open comments.

**Conclusion:** This Delphi study provides the first international consensus of 71 statements on return to sports, exercise, and physical activity following spinal fusion in AIS.

## Introduction

Adolescent idiopathic scoliosis (AIS) affects approximately 2-3% of the population and is characterised by a complex three-dimensional spinal deformity (1, 2). Typically, around 10% of individuals with AIS, with curves that measure above 50^0^, will go onto have spinal fusion, and biopsychosocial rehabilitation forms an essential part of recovery (3–7).

Following spinal fusion in AIS many individuals experience a reduction in muscle power, reduced range of motion and flexibility, decreased musculoskeletal function, and overall deconditioning (8, 9). Additionally, individuals have reported an increase in pain and fear causing between 28.0-36.6% of individuals to choose lower impact activities post-operatively (9). Following surgery 32.2% do not return to pre-operative activities (10). However, movement and activity post-operatively have been shown to reduce ongoing disability, dissatisfaction, societal and economic costs as well as the requirement for further operative intervention (11). Furthermore, psychological interventions delivered post-operatively improve pain, anxiety, quality of life, and satisfaction (12, 13).

Both physiotherapists and surgeons have a role in promoting post-operative return to sports, exercise, and physical activities (11, 14). Furthermore, physiotherapeutic interventions have been demonstrated as essential in promoting return to function in lumbar fusion (11). Pre-operative ambulation exercises in AIS have been shown to improve post-operative recovery (15). Immediate post-operative rehabilitation has been well documented in AIS such as strengthening, flexibility, and early day 1 mobilisation improving pain, recovery, and an earlier hospital discharge (16–19). A recent Delphi study explored surgeon consensus on post-operative care following posterior spinal fusion in AIS, but focused on interventions used in the acute phase during hospital stay rather than during outpatient rehabilitation (20). There is very little evidence for rehabilitation after an individual has been discharged from the acute setting (9, 11).

The International Society On Scoliosis Orthopaedic and Rehabilitation Treatment (SOSORT) supports the inclusion of the wider multidisciplinary team (MDT) in post-operative care (12). However, there remains a lack of consensus regarding which MDT members are necessary in the return to sports, exercise, and physical activity post-operatively (21, 22). At present most physiotherapeutic outpatient care remains surgically guided with variability worldwide seen between surgeons and between departments (21, 23). There is documented variability between physiotherapists regarding the reported rehabilitation interventions that are deemed to be most effective, alongside an array of biological, psychological, and sociological adjuncts to care (12, 24–28). Furthermore, there is evidence to suggest variability and a lack of consensus in care on a variety of issues such as rehabilitation milestones, post-operative protocols, and at what point it is necessary or safe to commence exercise and return to sports, exercise, or physical activity (9, 11, 23). This lack of consensus has resulted in a large spectrum of different rehabilitation approaches amongst those involved in the management of AIS, ranging from highly conservative immobilisation to early mobilisation and return to sports, exercise, and physical activity (23). This lack of clarity in rehabilitation has resulted in mixed expectations amongst medical professionals, patients, and caregivers, patients feeling fearful to move, participate in physical education classes at school, sports clubs, and ultimately lacking understanding regarding their care (10, 23).

Consequently, a study establishing consensus in post-operative rehabilitation and return to sports, exercise and physical activity is required. Return to sport can be influenced by a large number of factors such as parental influence, socioeconomic, and psychosocial variables (10). Furthermore, rehabilitation interventions might contain a spectrum of different biopsychosocial factors including restoring normal movement patterns and the promotion of self-management, education and advice, exercise, physical activity, or sports participation (7, 29, 30). Therefore, research that addresses this area must consider specific concepts such as the content of any rehabilitation, the philosophical approaches used and the milestones achieved for the longer-term outpatient physiotherapeutic interventions that promote a return to sports, exercise, and physical activity. Consideration of the role of the MDT will also be key in understanding post-operative rehabilitation.

### Aim

To determine global surgeon and physiotherapy expert consensus on the rehabilitation milestones and philosophical approaches taken in the return to sports, exercise and participation in physical activity for the postoperative care of those with AIS who have undergone spinal fusion surgery.

### Objectives

1. To determine surgeon and physiotherapy expert consensus on timelines and milestones in returning to sports, exercise and physical activity participation in AIS.
2. To determine expert consensus on the involvement of the MDT and philosophical approaches used for the postoperative rehabilitation interventions for AIS.

## Methods

### Study Design

This international three round electronic Delphi study was completed between 9^th^ February and 11^th^ November 2024. This study was performed according to a published open access protocol and is reported in accordance with the Conducting and REporting Delphi Studies (CREDES) guidelines (31–33).

### Sample and expert eligibility

This Delphi survey was distributed to a snowball sample of surgeon and physiotherapy experts working internationally within the management of AIS.

A sample of surgeon and physiotherapy experts from two populations were recruited (34):

1. Physiotherapy or surgeon academics with ≥2 publications on AIS, paediatric spinal pain, or adolescent spinal pain in peer reviewed journals within the last 10 years.
2. Clinical surgeon or physiotherapy experts with a caseload of ≥20% spinal deformity practice specifically in AIS (either conservative or operative) per month.

### Recruitment

Recruitment took place from 9^th^ February 2024 to 11^th^ June 2024. Participants were contacted via scoliosis professional network calls, publicly available non-NHS emails, and social media snowballing (35). The survey was sent electronically to individuals from a range of diverse backgrounds and a variety of geographical locations (31, 36). Professional networks included the British Scoliosis Society, Scoliosis Association UK, Scoliosis Support and Research, Scoliosis Australia, Straight Caribbean Spine Foundation, Spine & Scoliosis Research Associates Australia, Scoliosis Research Society, National Scoliosis Foundation, Bundesverband Skoliose-Selbsthilfe, Vereniging van Scoliosepatiënten, Verein Scoliose Schweiz, Setting Scoliosis Straight, International Society On Scoliosis Orthopaedic and Rehabilitation Treatment, and AO Spine.

### Sample size

In order to achieve consensus, a minimum of 27 experts were required to complete all three rounds (37–41). Due to an estimated response rate of 70% this study aimed to recruit a minimum of 40 consenting experts (20 surgeon and 20 physiotherapy experts) (37–40, 42, 43). To help reduce risk of drop-out, experts chosen were willing individuals who have interest and subject specific knowledge rather than enticement through monetary reward (36, 44–47).

### Procedure

This Delphi study consisted of three iterative rounds. Figure 1 shows a summary of the Delphi rounds.

**Figure 1.**
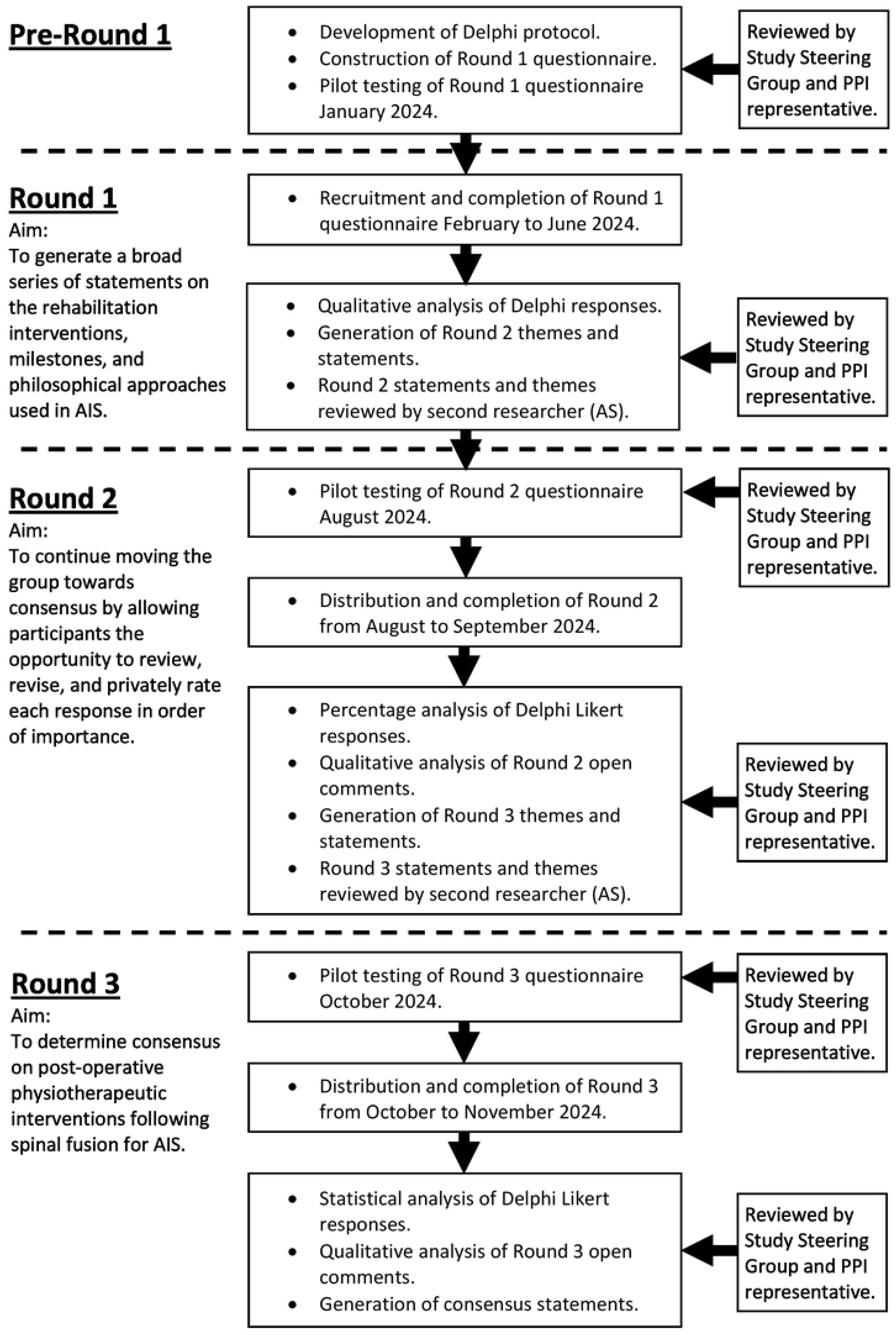

### Pilot Testing

Round 1 was piloted with 1 physiotherapist and 1 surgeon from a tertiary hospital specialising in the care of those with AIS, both participants who completed the pilot testing of rounds were not included in the main study (36). Piloting of the round 1 survey took place in January 2024 prior to distribution of the round 1 survey. All 3 rounds were piloted with the same two individuals (36).

### Round 1

The aim of round 1 was to generate a broad series of statements on the rehabilitation interventions, milestones, and philosophical approaches used in AIS. The study steering group (AS, NH, AG, AR) and PPI representative (ER) reviewed the questionnaire pre- and post-piloting of the survey (48, 49). During round 1 individuals first completed an online information and consent form. Following consent and questions on demographic data, participants were asked a series of open-ended questions and asked to anonymously express their views with freedom in responses (50, 51). Data were also gathered on the individual’s geographical location, percentage of practice involving spinal deformity, and nature of either clinical or academic work.

Inductive content analysis was used to identify themes, patterns, or concepts (52, 53). All results were verified by a second researcher (AS) and reviewed by the study steering group to ensure that data was represented fairly (51). Dissonance was highlighted for consideration in round two in an attempt to move towards whole group consensus (54). Data on participant characteristics including location and volume or nature of work was also tabulated to aid the analysis of results and illustrate any potential confounders or mediators in results analysis. Consensus during round 1 was defined as agreement >1 participant for responses given. A summary of qualitative findings were included at the start of Round 2 data collection following discussion with the study steering group (36).

### Round 2

The aim of round 2 was to continue moving the group towards consensus by allowing participants the opportunity to review, revise, and privately rate each response in order of importance (35). All participants from round 1 were invited to take part in round 2, including those who did not complete round one, to help reduce the risk of false consensus (55). Ideas and statements generated during round one with agreement >1 were re-distributed during round two with any dissonance between the physiotherapy and surgeon experts summarised. During round two participants were invited to rate their agreement with statements using a 5-point Likert scale, 1 = strongly disagree, 5 = strongly agree. Participants were also given the opportunity to offer their opinion in an open text box in response to themes and statements redistributed.

Likert rated numerical data from round two was quantitatively analysed using descriptive and inferential statistics (51). Statements rated with either agree or strongly agree were used to create a percentage to evaluate consensus amongst experts for each statement (48). Central tendencies of rated items (median) and dispersion (interquartile range) were calculated summarised at the start of round three (51, 54). Any open-ended responses that had agreement across >1 participant were grouped and summarised for inclusion in round three once verified by the second researcher (AS) (31, 51, 54). Consensus during round two was determined when >75% participants within the group agreed with a statement (56).

### Round 3

The aim of round three was to determine consensus on post-operative physiotherapeutic interventions following spinal fusion for AIS. Participants were provided with a summary of the results from round two with areas of dissonance identified. Summary data including descriptive and inferential statistics, dissonance, and stability (49, 51) were visible to all consenting participants. Those who have withdrawn, regardless of their central tendency or dispersion, were excluded (50). Participants were asked to rate their agreement with the statements that achieved >75% agreement during round two, using the same 5-point Likert scale but with an additional opportunity for open comments.

Round 3 rated numerical data was quantitatively analysed in the fashion already described for round 2 with statements >75% agreement put forwards for further statistical analysis. Round three statements were statistically analysed using Kendall’s coefficient of concordance (57). It was determined that consensus would be achieved when Kendall’s coefficient ≥0.7 (58–61). Stability between round two and three was calculated using a Wilcoxon rank-sum test with significance at P<0.05 (49, 54, 62, 63). Consistency and stability between responses between Rounds was considered a necessary criterion for stopping (54).Data was tabulated and presented summarising results and consensus on post-operative interventions for return to sports, exercise, and physical activity. During round 3 there was a final opportunity for open comments. Comments made >1 participant were formed into statements. However, these final statements were not rated by all participants on the Likert scale and therefore have not been subject to statistical testing from Kendall’s W or Wilcoxon sum rank test for stability. These additional statements included in the final consensus have been recorded in a separate table to provide clarity for the reader.

### Definition of consensus, agreement and stability

During each round, consensus, agreement and stability were assessed (Table 1).

**Table 1:** Definitions used.

| Item | Definition |
| --- | --- |
| **Stability across all rounds** | Consistency of responses and presence of consensus amongst the group for each statement between Delphi rounds two and three using a Wilcoxon sum-rank test (48, 54, 63). |
| **Agreement** | The level of agreement between expert participants as demonstrated by the percentage, thereby enabling us to predict the rating of another expert (54). |
| **Consensus Round 1** | Agreement >1 participant for statements made. |
| **Consensus Round 2** | Level of agreement $\geq 75\%$ between participants for each statement. |
| **Consensus Round 3** | Kendall's coefficient of concordance = 0.7 (58-61). |

### Data management

All rounds of both the pilot and the final Delphi were completed using Research Electronic Data Capture (REDCap) software for electronically distributing and collecting survey data (64, 65).

The three rounds of the Delphi study distributed to surgeon and physiotherapy experts is summarised in Figure 1. Participants did not meet directly during this study but instead were sent online questionnaires seeking views on the topic of interest (50). The survey and survey data was distributed, collected, stored and analysed electronically using REDCap (65). REDCap is a secure online software that is designed for creating and managing online surveys (66). All data is securely stored using REDCap on a password-protected computer with members of the research team only having access to the data. Following completion of the study, data will be securely stored within the University of Birmingham, UK for 10 years following which it will be safely destroyed in accordance with the University of Birmingham guidelines.

### Study steering group

Co-authors (AS, NH, AG, AR) constitute the study steering group, comprising methodological, academic expertise and clinical expertise in physiotherapy and spinal surgery. The group met to discuss the results analysis from each round and iteration in the subsequent round and to provide feedback and critical insights on the progress of the project.

### Patient and public involvement (PPI)

A PPI representative (ER) has been involved from study conceptualisation until final dissemination. Both clinicians and academics working within the field of AIS were involved in making research decisions and giving feedback regarding methodology and results synthesis at all stages of the process. The Guidance for Reporting Involvement of Patients and the Public short form checklist (GRIPP2-SF) has been used to promote PPI reporting (67) (Supplementary File 1).

## Results

Fifty-three eligible experts consented to participate in this study (27 surgeons and 26 physiotherapists) (Table 2).

**Table 2:**
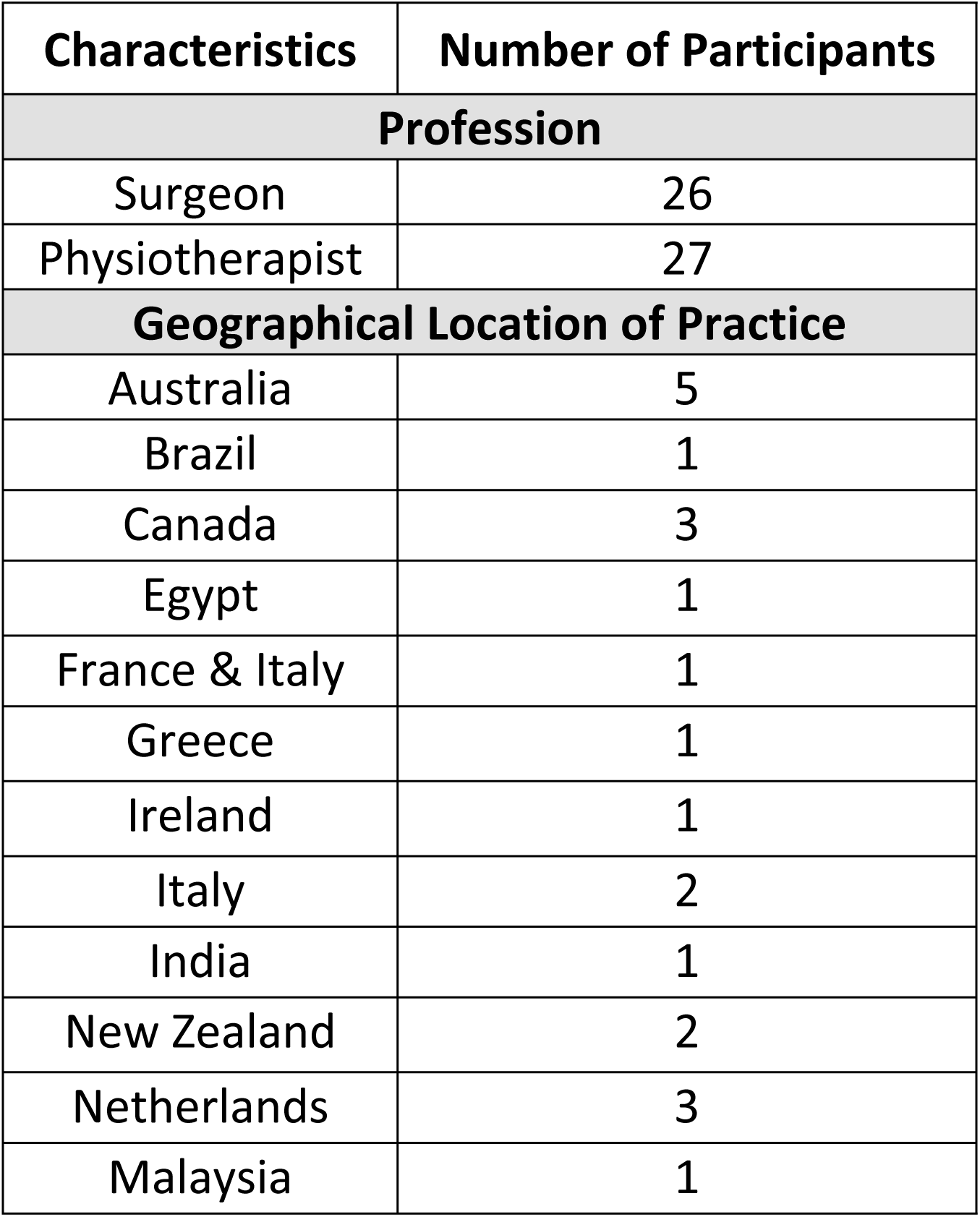
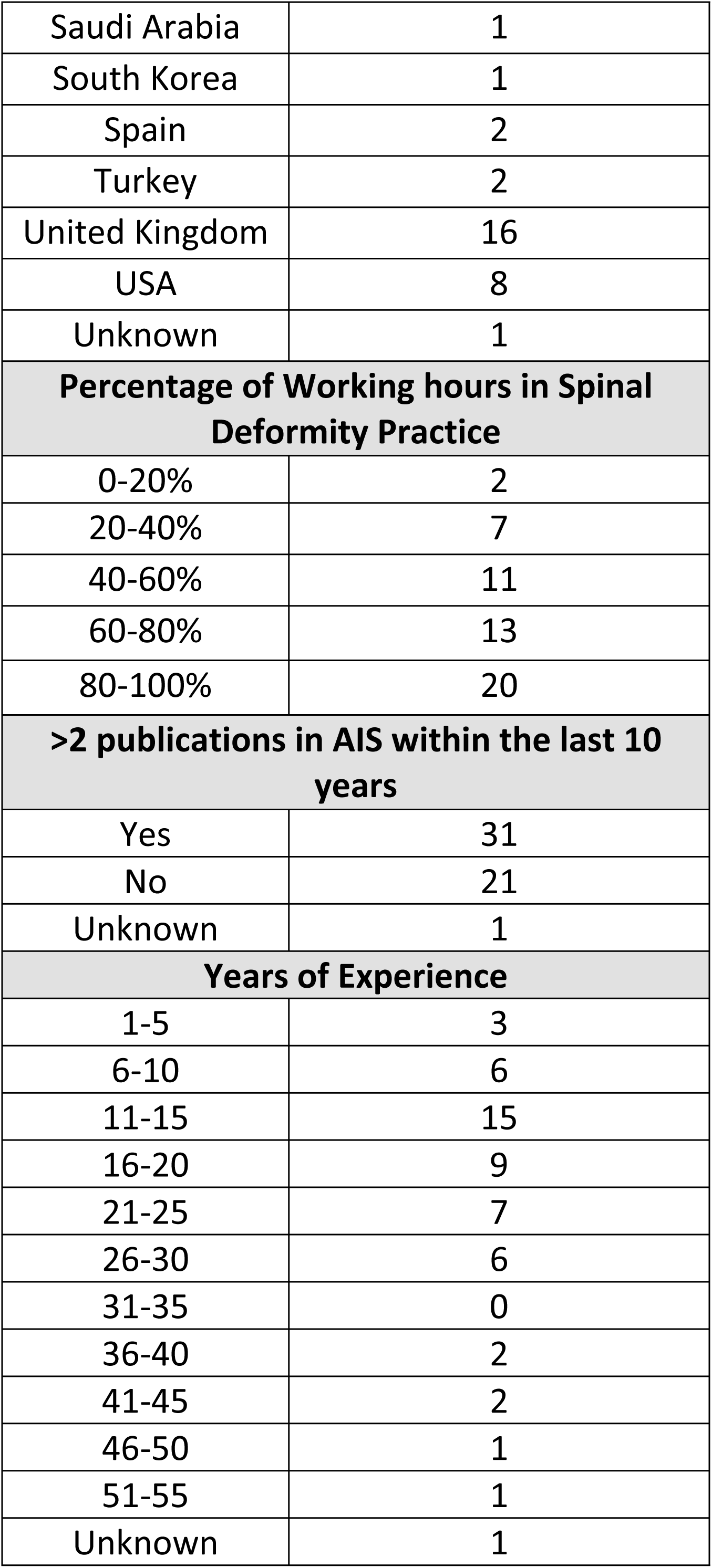
Demographic Data.

| Characteristics | Number of Participants |
| --- | --- |
| **Profession** |  |
| Surgeon | 26 |
| Physiotherapist | 27 |
| **Geographical Location of Practice** |  |
| Australia | 5 |
| Brazil | 1 |
| Canada | 3 |
| Egypt | 1 |
| France & Italy | 1 |
| Greece | 1 |
| Ireland | 1 |
| Italy | 2 |
| India | 1 |
| New Zealand | 2 |
| Netherlands | 3 |
| Malaysia | 1 |
| Saudi Arabia | 1 |
| South Korea | 1 |
| Spain | 2 |
| Turkey | 2 |
| United Kingdom | 16 |
| USA | 8 |
| Unknown | 1 |
| **Percentage of Working hours in Spinal Deformity Practice** |  |
| 0-20% | 2 |
| 20-40% | 7 |
| 40-60% | 11 |
| 60-80% | 13 |
| 80-100% | 20 |
| **>2 publications in AIS within the last 10 years** |  |
| Yes | 31 |
| No | 21 |
| Unknown | 1 |
| **Years of Experience** |  |
| 1-5 | 3 |
| 6-10 | 6 |
| 11-15 | 15 |
| 16-20 | 9 |
| 21-25 | 7 |
| 26-30 | 6 |
| 31-35 | 0 |
| 36-40 | 2 |
| 41-45 | 2 |
| 46-50 | 1 |
| 51-55 | 1 |
| Unknown | 1 |

### Participant Demographics

Demographic data regarding profession, geographical location, experience, and practice is summarised (Table 2). There were 26 surgeons (49%) and 27 physiotherapists (51%). Experts were from 18 different countries (1 unknown) spread across all 5 continents. Fifty-one (96%) participants worked >20% of their hours in spinal deformity practice, 2 participants (4%) worked less than 20% of their hours in spinal deformity practice. However, these 2 participants were both eligible to participate in the e-Delphi for having >2 publications within last 10 years.

### Round 1

Fifty-three participants were included in the study because they were both eligible and gave consent (from the initial 60 participants who gave consent). Forty-one out of fifty-three experts (77%) commenced Round 1 and eighteen experts completed all questions in Round 1 (Figure 2). During Round 1, 85 statements were generated with agreement >1 participant. These 85 statements were grouped under 19 themes (Appendix 1) and redistributed during round 2 for Likert rating.

**Figure 2.**
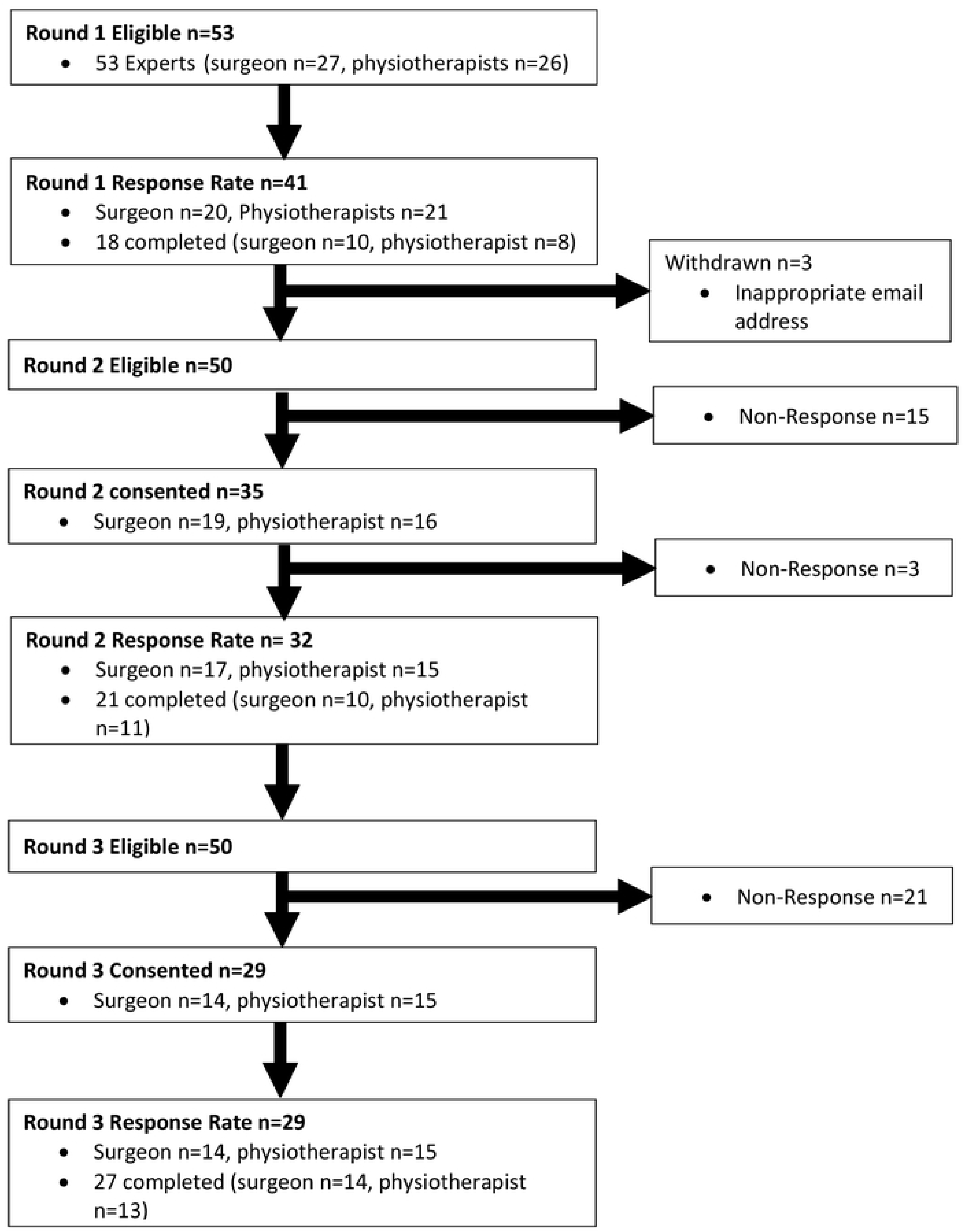

### Round 2

Thirty-five of the fifty eligible experts consented to complete Round 2 and 32 participants commenced Round 2. Of these twenty-one completed Round 2. During Round 2 56 out of 85 statements reached > 75% agreement on the 5-point Likert scale (5 = Strongly Agree or 4 = Agree) (Appendix 2). Of the 56 statements agreed in round 2 seven explored multiple concepts and each of these seven statements were split into two statements making a total of 63 statements put forwards into round 3.

Despite multiple data checks there were four statements progressed to Round 3 with <75% agreement due to a processing error. However, this was visible to all participants during round three with bar charts showing levels of agreement. During Round 3 two of these four statements (Strengthening Exercises and Cardiovascular Fitness modalities) were then included in the final consensus due to achieving >75% agreement in Round 3. Meanwhile, two statements regarding a cognitive behavioural approach and commencing water-based exercise / rehabilitation or hydrotherapy as soon as the wounds are healed (approx. 2 weeks post-op) both scored <75% agreement therefore were removed from the final consensus. These 4 statements are highlighted in Appendix 2.

Open comments from Round 2 were analysed and where there was consensus (frequency >1 participant) on a comment or theme an additional statement was generated (Appendix 2). Interestingly, Round 2 open comments generated 2 statements regarding hydrotherapy or water-based exercise highlighting dissonance on opinion of the role of water or hydrotherapy (Appendix 2). In total there were 22 additional statements generated from Round 2 open comments. In total 85 statements were put forwards for rating in round 3. These statements were arranged into 10 themes regarding different aspects of care.

### Round 3

Twenty-nine participants consented to Round 3 (14 surgeons and 15 physiotherapists). Of these 29 participants, 27 participants answered all questions and completed to the end of Round 3 (surgeon 14, physiotherapist 13). From the 85 statements distributed at the start of Round 3 (56 round 1 statements and 29 additional statements generated from the round 2 comments) 67 out of 85 statements achieved >75% agreement (5 = Strongly Agree or 4 = Agree). For these 67 statements Kendall’s Coefficient of Concordance (W) was calculated at 0.5 moderate agreement (Table 3) (58). As part of exploring dissonance the differences in consensus between surgeons and physiotherapists were evaluated (Table 3). Both groups demonstrated significance in the results and consensus generated, but the surgeons had greater agreement (W=0.5, p<0.001) compared with Physiotherapists (W=0.2, P<0.001).

**Table 3:** Inter-rater Kendall’s coefficient of concordance for round 3 statements.

| Round 3 Statements | All 29 Participants |  | Surgeons |  | Physiotherapists |  |
| --- | --- | --- | --- | --- | --- | --- |
|  | W | P | W | P | W | P |
| All 85 Round 3 statements rated Strongly Agree or Agree | 0.442 | <0.001 | 0.456 | <0.001 | 0.465 | <0.001 |
| Final 66 Round 3 statements that had >75% Strongly agree or agree | 0.469 | <0.001 | 0.473 | <0.001 | 0.191 | <0.001 |

From the 85 statements that were generated at the end of round 2 66 statements achieved >75% agreement in Round 3. The final 71 consensus statements were from 66 round 3 Likert rating and 5 statements were generated from Round 3 open comments. The Wilcoxon sum rank test demonstrated stability with P>0.05 across all 63 statements.

During the Delphi process the number of consensus statements put forward into round 2 were 85 statements (Appendix 1). During round 2 this was reduced to 56 with 22 round 2 comment statements. A total of 85 statements were put forwards into round 3 as 7 of the 56 original statements were split into 63 due to multiple concepts being explored. During round 3 this reduced to 66 statements (Table 4) with five additional round 3 comments (Table 5) making a total of 71 statements. Within the final 71 consensus statements 14 out of the 22 round 2 additional comments resulted achieved consensus (Figure 3).

**Table 4:**
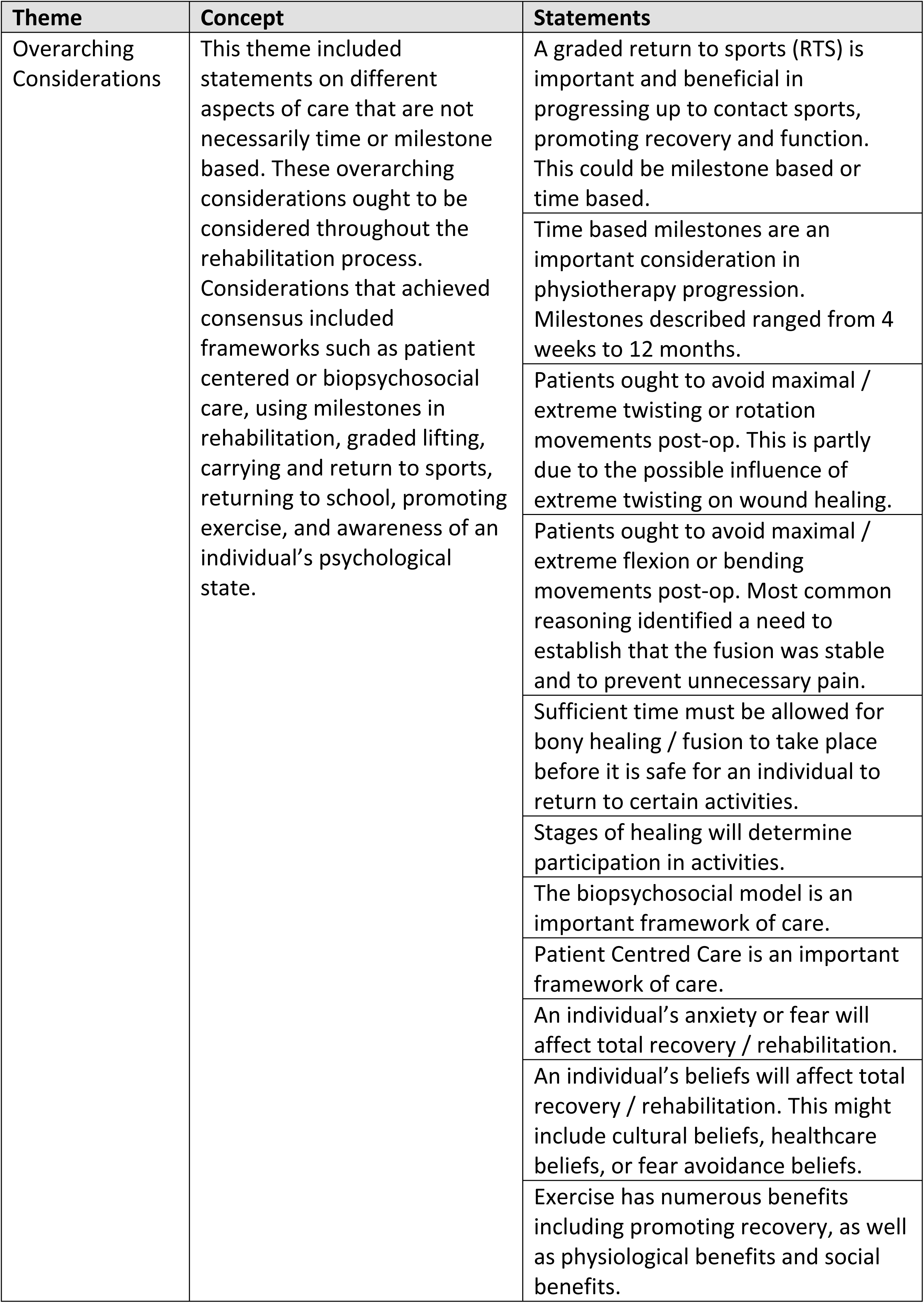
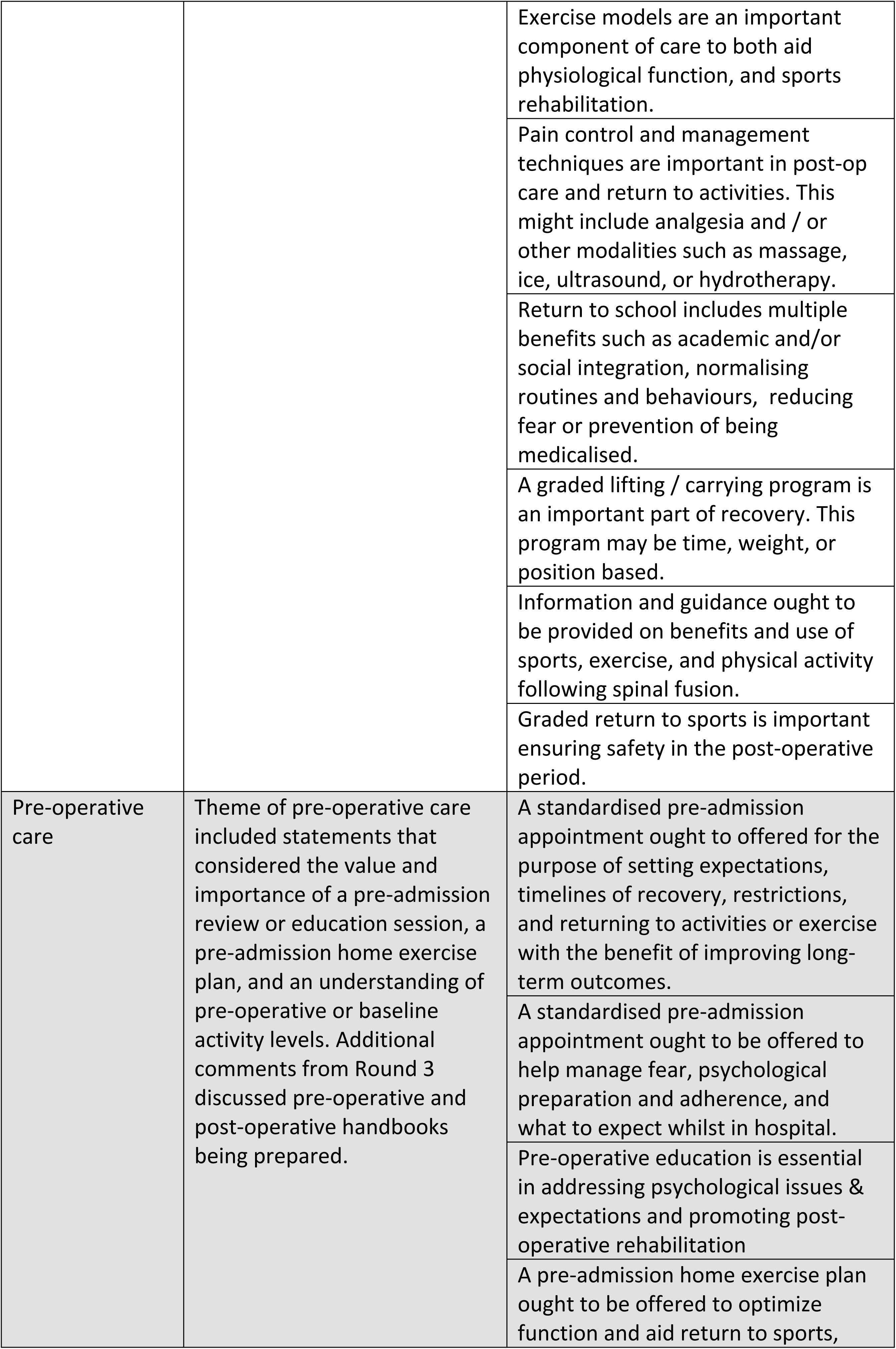
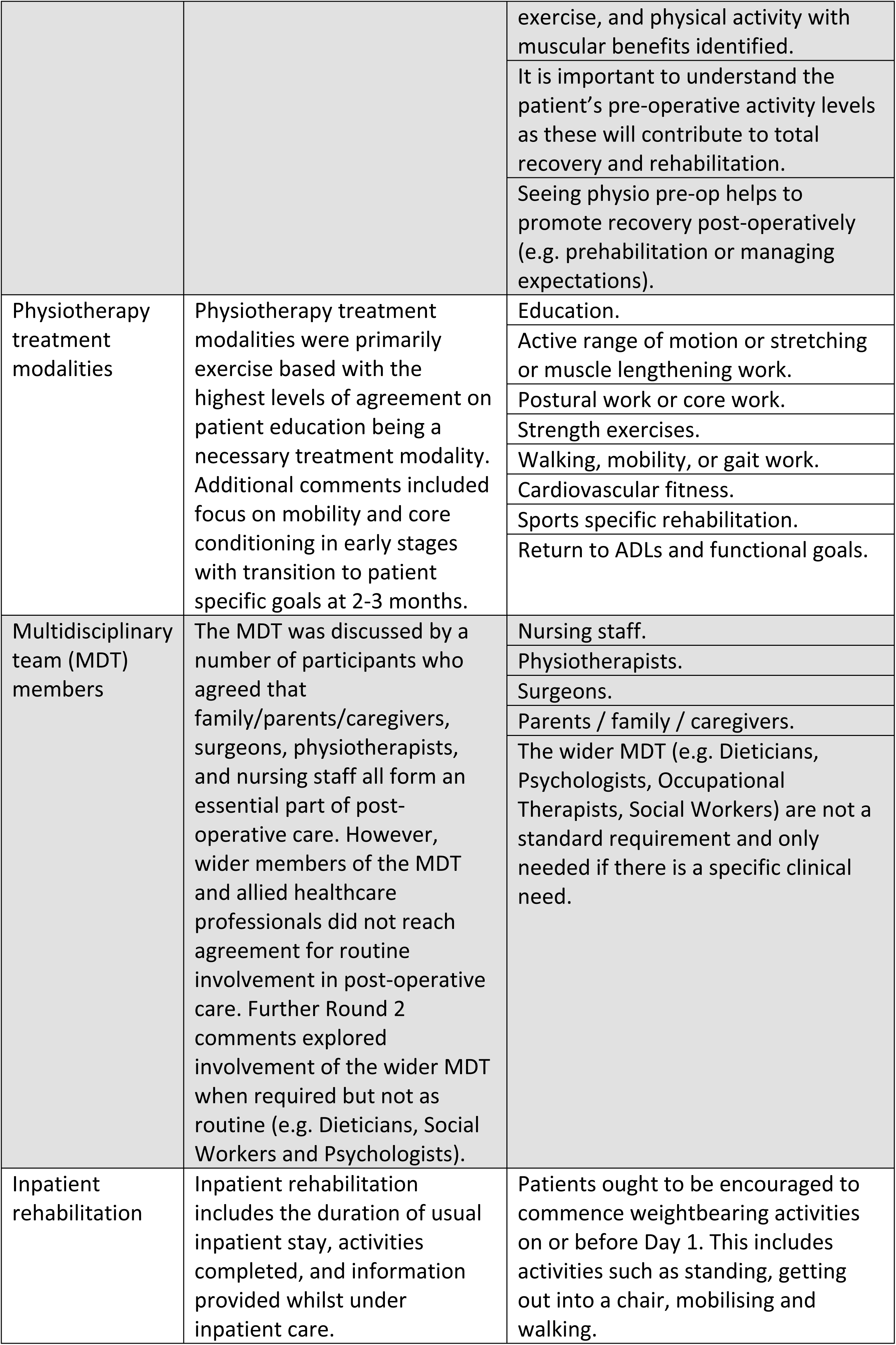
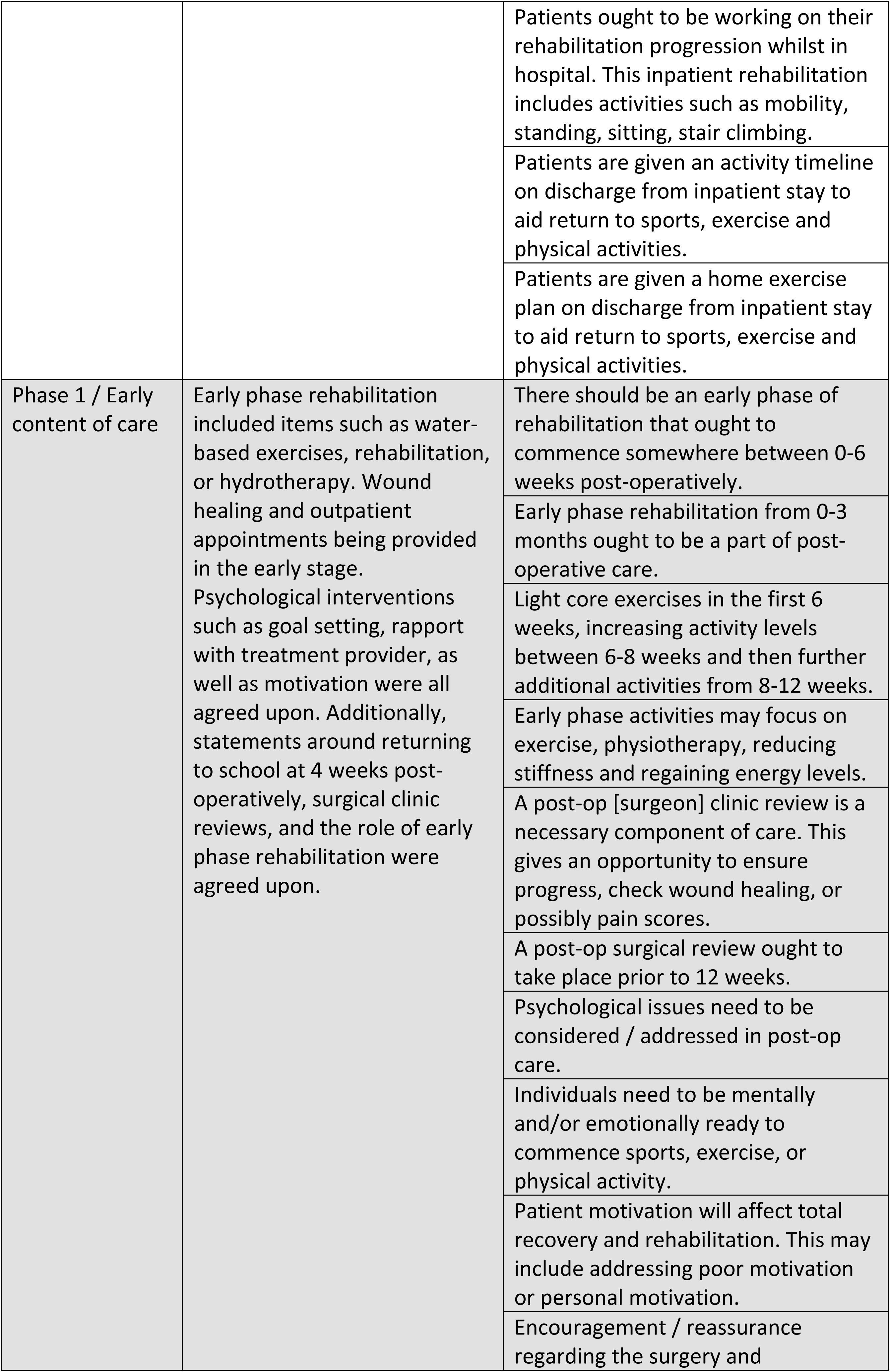
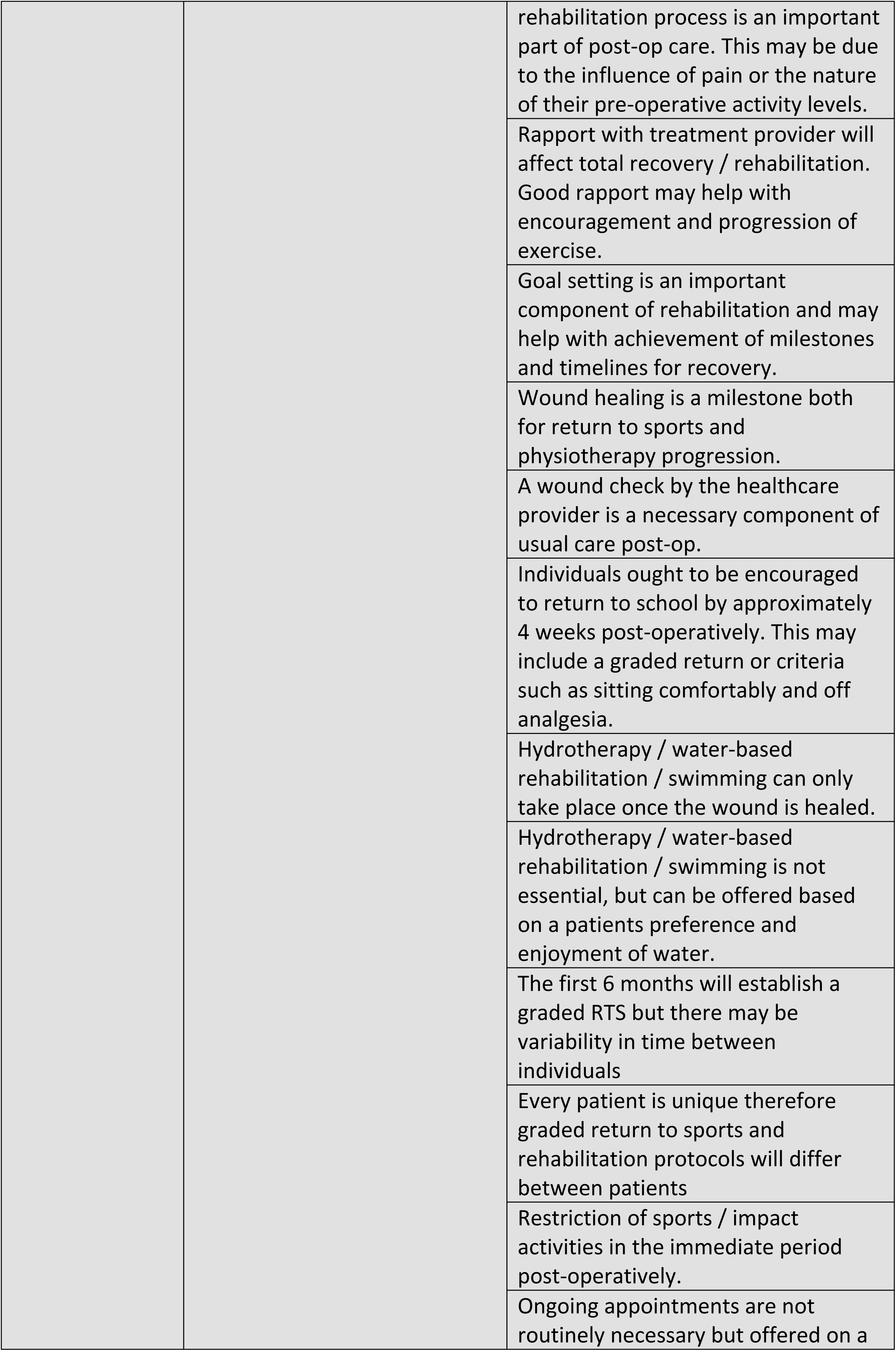
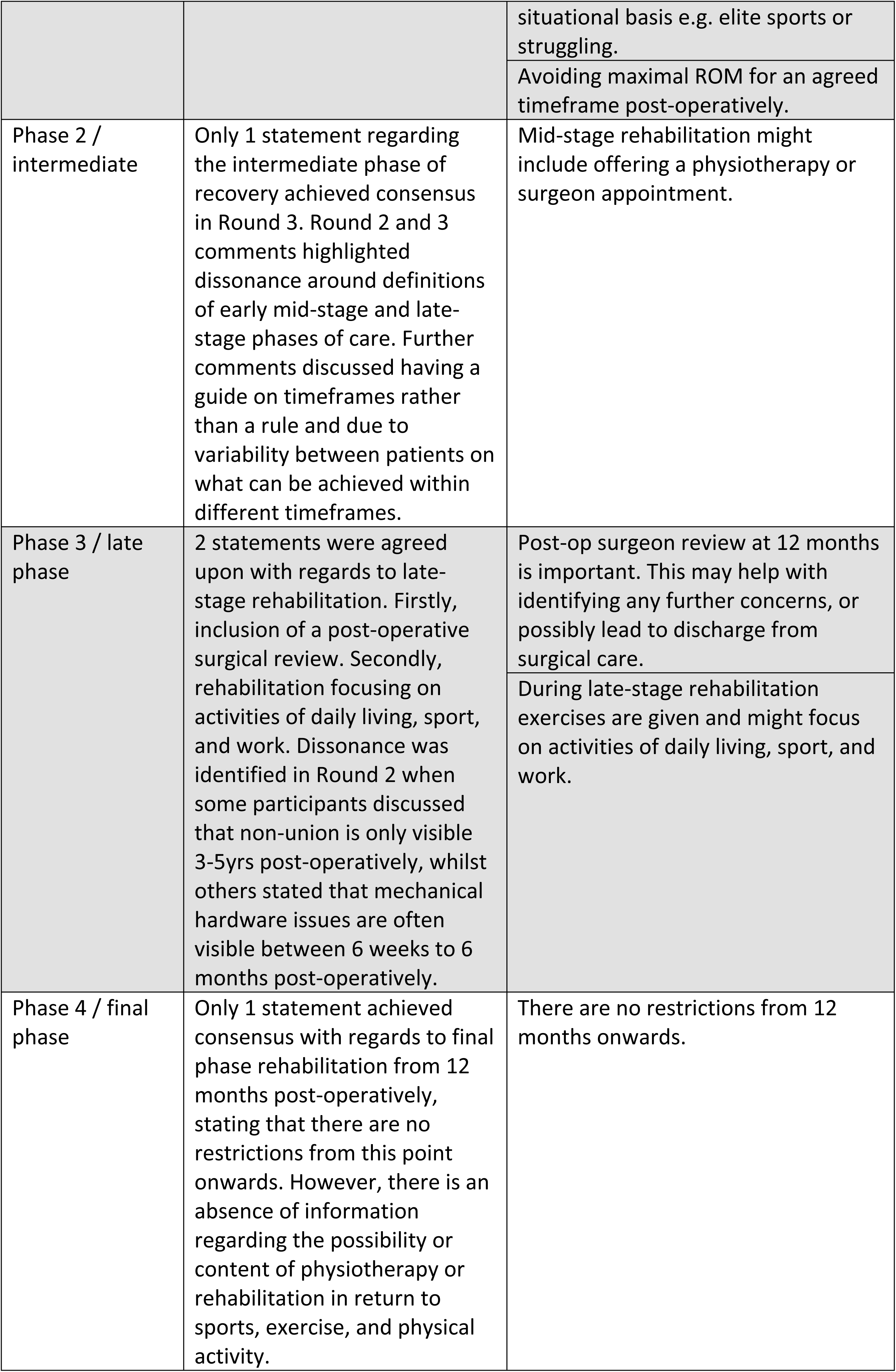
Consensus Themes, Concepts, and Statements.

**Table 5:** Statements generated from Round 3 further comments.

| Statement | Supporting Comments |
| --- | --- |
| Scoliosis specific exercises (SSE) might be appropriate in some cases | "Scoliosis Specific Exercises may be needed but not in everyone". |
|  | "Re-education on postural and muscular balance and physiological breathing function and activities of daily living (ADL) activities with the guidance of educated possibly physiotherapeutic scoliosis specific exercises certified physiotherapists". |
| Post-operative physiotherapy ought to be provided based on need rather than given to every patient | "Similarly, post-op physio needs assessing rather than just something that can be done in everyone as this is not realistic and would waste valuable resource". |
|  | "Rehabilitation from physiotherapy once discharged home can be done on a patient need basis. Need for input can be identified in clinic on medical review". |
|  | "Not everyone needs a rehab programme". |
|  | "Post-surgery we offer specific rehabilitation on patient need basis". |
| Timeframes for milestones and rehabilitation will vary between patients and are dependent on a variety of unique factors | "Milestones described ranged from 4 weeks to 12 months. Milestones vary from person to person. It is not possible to set an exact number on the milestones". |
|  | "I don't think trying to define mid-stage, intermediate phase, or late-stage rehab is really that helpful. Each patient is going to |
|  | progress through their rehab based on several unique individual variables and what their personal physical goals are". |
|  | "This is a vague period that is going to be highly variable between patients". |
| Respiratory exercises might be appropriate in some cases as part of rehabilitation | "Respiratory exercises or incentive spirometry is the milestone of healing". |
|  | "The patients need respiratory exercises". |
| Specialist members of the multidisciplinary team can perform post-operative reviews (e.g. physios or nurses) | "Post-operative reviews are important but don't necessarily need to all be done by the surgeon as a well-trained nurse practitioner, physiotherapist, physician or other member of the team would likely be able to address most patient concerns/questions". |
|  | "Our service has a 6-week review post op but this is carried out by a specialist nurse or physiotherapist". |

**Figure 3.**
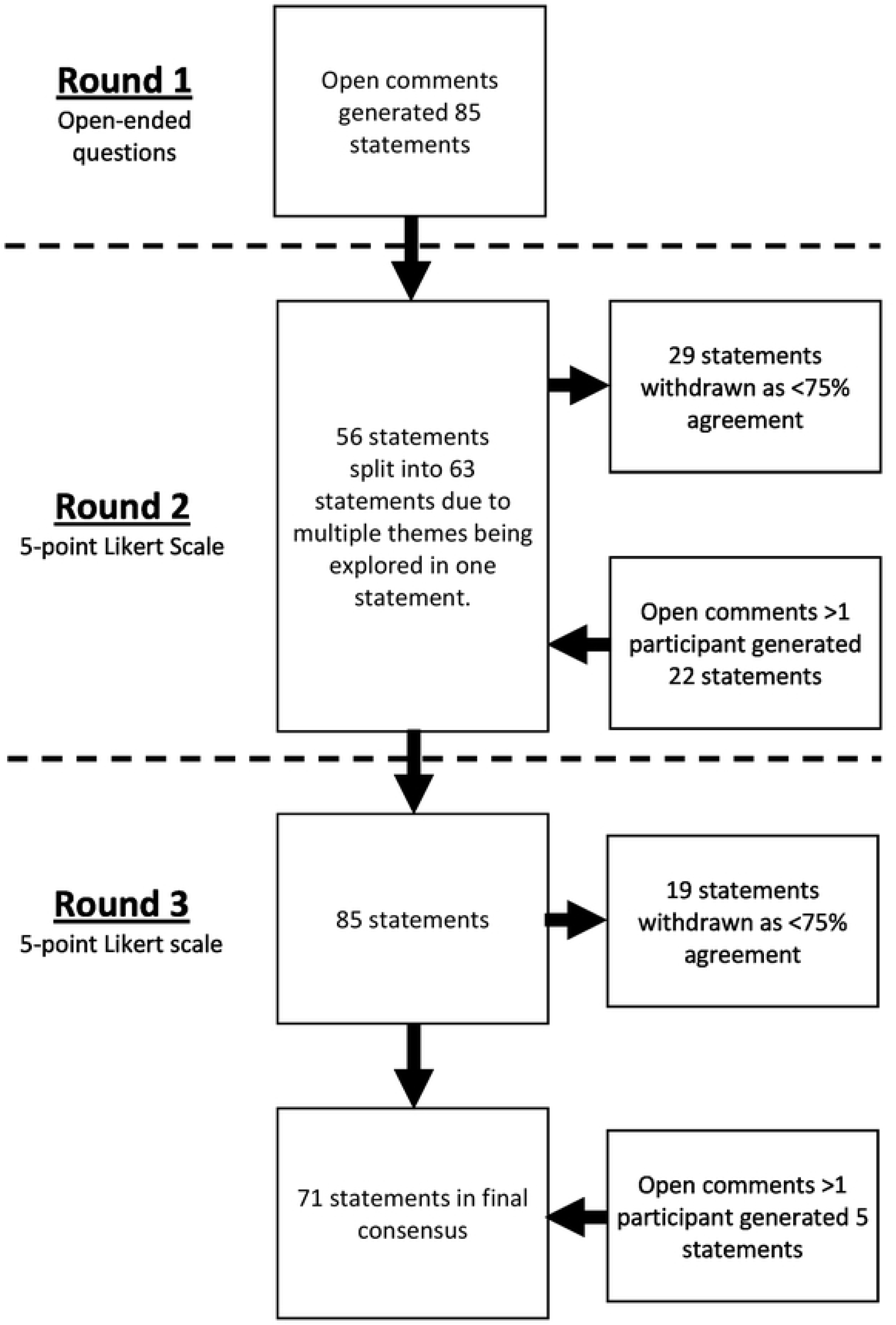

Statements that achieved consensus during round 3 comprised of 9 themes: (1) Overarching considerations, (2) Pre-operative Care, (3) Physiotherapy Treatment Modalities, (4) MDT involvement, (5) Inpatient Rehabilitation, (6) Early / Phase 1 Care, (7) Intermediate / Phase 2 Care, (8) Late / Phase 3, and (9) Final / Phase 4. The theme of overarching considerations and physiotherapy treatment modalities are not restricted to a certain timepoint / stage of the process from decision to undergo scoliosis correction via spinal fusion, but rather are components of care that may be used at any stage in the process. All statements regarding components of care that achieved consensus are listed in Table 4 related to their respective themes. Figure 4 demonstrates the international consensus in a form that may provide appropriate guidance for healthcare professionals.

**Figure 4.**
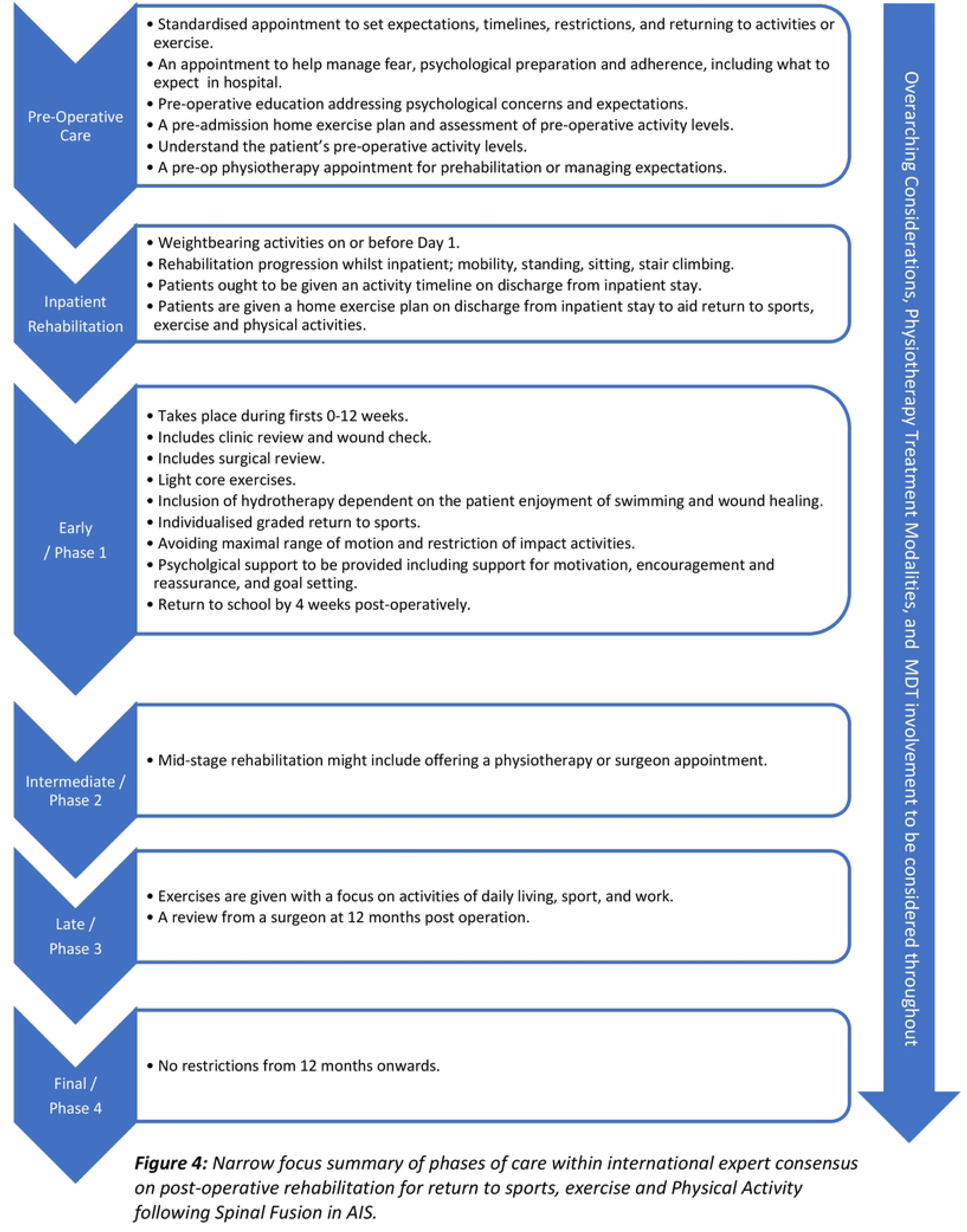

During Round 3 there was a final opportunity for open comments. The qualitative data captured some further issues that are still important for participants at the end of the Delphi process. A further 5 statements were generated from these open comments with agreement >1 participant (Table 5). At the end of the consensus processes ongoing issues discussed in comments included scoliosis specific exercises, post-operative rehabilitation, timeframes, respiratory exercises, and involvement of specialist members of the MDT in post-operative reviews.

## Discussion

This study was the first to establish a consensus on timelines, milestones, and content of rehabilitation for return to sports, exercise, and physical activity following spinal fusion in AIS. This Delphi generated a series of consensus statements that relate to nine major themes as well as the role of the MDT and philosophical approaches to care.

### Expert consensus on timelines and milestones

A large number of statements were generated supporting rehabilitation in the initial and early phases of care. However, the Intermediate Phase 2, Late Phase 3, and Final Phase 4 stages of rehabilitation in return to sports, exercise, and physical activity remain underrepresented with 1 statement for both Intermediate and Final and 2 statements for Late phase rehabilitation. This demonstrates dissonance in relation to earlier statements regarding the need for a graded return to sports and sports specific rehabilitation. The 2016 consensus statement on return to sport describes a continuum paralleled with recovery and rehabilitation with three phases (1) return to participation, (2) return to sport, and (3) return to performance (68). However, beyond Early Phase 1 rehabilitation this Delphi consensus lacks further detail with regards to return to sport and performance, thereby warranting future exploration in late-stage sports, exercise, and physical activity post-operatively. Graded return to sports is a complex multifactorial process and has been recognised an a detailed part of rehabilitation (69). There is limited evidence surrounding physiotherapists or other medical or healthcare professionals’ confidence or knowledge regarding administration of a return to sport program, particularly within spinal pain. However, one study concluded that a lack of knowledge and training was a barrier to implementing psychological strategies in return to sports rehabilitation (70). Further, exploration of physiotherapists’ confidence, knowledge, content, and application of return to sports programs is necessary, particularly following spinal fusion in AIS. Furthermore, this Delphi highlighted vagueness with regards to timeframes for return to sports, exercise, and physical activity. This is likely to make it more acceptable to professionals involved in the rehabilitation and return to sports, exercise, and physical activities in allowing individual tailoring of guidance in accordance with the specific and complex biopsychosocial needs of the patient (71).

### MDT involvement

The role of the MDT was a key objective of this study as SOSORT recommendations support the involvement of the wider MDT (12). However, which members of the MDT ought to routinely be involved in care, particularly when supporting post-operative return to sports, exercise, and physical activity was not defined (21, 22). Experts highlighted a wide variety of MDT members who may not be routinely included in post-operative rehabilitation but have a valuable role in supporting care if there is a specific clinical need. This is the first study to explore who from the MDT ought to be involved in post-operative return to sports, exercise, and physical activity in AIS. Other literature has evaluated the benefits of the MDT in contexts such as cancer care and primary care with evidence demonstrating an improved quality of care (72–74). However, further work is needed exploring the involvement and benefits of the MDT in AIS. Parents, family, or caregivers were also determined as having a necessary involvement in post-operative rehabilitation. Although not a part of the professional MDT literature shows that parents experience substantial emotional burden regarding their child’s future, work opportunities, ongoing back problems and significant concern around consenting for spinal fusion surgery (75, 76). Furthermore, parents may be more worried than their child following a diagnosis of AIS and literature shows that a parent’s response can either promote or confound a child’s health outcomes (75, 77). Therefore, this consensus supporting the involvement of parents, family and caregivers is in keeping with the literature demonstrating the value and benefit of parental information and support on a child’s post-operative outcomes (76, 77).

### Philosophical approaches to rehabilitation

This study set out to explore philosophical approaches underpinning post-operative rehabilitation. Both the biopsychosocial model and patient centered care were agreed upon as necessary frameworks providing a philosophy of care that guide the application of medical knowledge based on the needs and of and shared decision making with the patient (78, 79). Within this Delphi consensus are a series of statements on overarching considerations that ought to be considered at each stage of rehabilitation and can be selected based on patient needs and choice. Additionally, there are several statements that refer to the possible use of an intervention at a specific stage based on the patients needs. Although, the biopsychosocial model has been criticized for its vagueness, it allows clinicians to broaden their gaze to a variety of factors that may influence return to sports, exercise and physical activity (78). Together both philosophical approaches allow clinicians to select and tailor appropriate interventions based on patient need and choice rather than a strict prescriptive guideline, improving application and outcomes of rehabilitation (79, 80).

### Strength of agreement in consensus

Study findings demonstrated moderate agreement (W=0.5, p<0.05) (58). Although, we set out to achieve strong agreement (W=0.7) the significance of the p value demonstrates concordance between experts (58, 81). The definition of consensus is contested in the literature and has not yet been established (54). However, level of agreement between experts is more easily defined with many studies using percentage agreement, particularly when Likert scales have been used (54). Our study first evaluated percentage agreement with consensus >75% (Strongly Agree or Agree), followed by Kendall’s coefficient of concordance demonstrating both strength of agreement and significance (56). It was decided in discussion with the study steering group that the presence of moderate agreement (W=0.5) and significance in results (p<0.05) provided an appropriate stopping criterion and a fourth round was not required (54). Furthermore, the challenges to a fourth round include participant attrition resulting in overestimation of consensus, particularly if participants with minority perspectives drop out (82, 83). Further research may usefully explore agreement distribution and has the potential to offer new perspectives, explore the multifactorial nature of post-operative rehabilitation, or offer a deeper understanding of factors influencing agreement amongst experts (84).

### Stability of consensus statements

For all statements stability was calculated between round 2 and round 3. The stability demonstrated no significant changes and experts were consistent in their views (p>0.05) (85). Furthermore, when performing multiple tests it is important to consider the possibility of Type 1 error and therefore need for a Bonferroni correction (85). In our study all statements were not significant therefore the addition of a Bonferroni correction did not change the results, for the sixty-three statements that were rated on the Likert in both Round 2 and 3 (p=0.0008), for the fifty-three statements that were both rated on the Round 2 and 3 Likert and had >75% agreement (p=0.001). Therefore, the Bonferroni correction was not necessary as all values were p>0.05 (not statistically significant).

### Consensus on the role of scoliosis specific exercises

Scoliosis Specific exercises (SSE) including Schroth are tailored exercises focusing on the three-dimensional treatment of AIS but realigning the spine, rib cage, shoulders, and pelvis (86). There are a variety of different schools that teach exercises that come under the definition of SSE and are recommended by SOSORT (12, 86). The effectiveness of SSE remains debated with one review reporting a short-term improvement in Cobb angle, trunk rotation, and quality of life (87), although the improvement in Cobb angle did not exceed the minimum clinically important difference (87). Meanwhile, another review concluded the evidence is insufficient to confirm the benefits of one specific physiotherapy intervention over another (88). The opportunity for further comments in this Delphi revealed that there is still debate amongst experts regarding SSE, with dissonance between participants convicted in their views. A statement around the use of SSE was generated from Round 1 and participants had the opportunity to vote on this during Round 2 (Appendix 1). During Round 2 and 3 Likert voting SSE was dropped due to a lack of consensus but re-introduced with open comments >1 participant (Table 5). This dissonance is perhaps reflective of the wider debate in the literature regarding the role of SSE (88). Furthermore, in post-operative spinal fusion, the Cobb angle has been objectively reduced through mechanical instrumentation (89). Therefore, unless there is presence of scoliotic curves outside the area of instrumentation, the requirement of SSE to reduce the size of a spinal deformity is no longer present (90). Consequently, the justification for the use of SSE post-operatively is reduced to only operated patients with pain to increase function as stated in SOSORT guideline (12, 91). However, further exploration of the statement generated by open comments is required to understand rationale for comments and application (84).

### Consensus on role of respiratory exercises

Spinal fusion in the treatment of scoliosis is known to have varying effects on respiratory function due to the effect on the chest wall, particularly with an anterior approach or thoracotomy (92). Posterior spinal fusion is thought to be less deleterious on post-operative respiratory function, with debate in the literature whether posterior fusion increases or maintains respiratory function (92–94). Statements around respiratory care became another example of dissonance with statements being regarding the role of respiratory exercises being included during round 2 alongside contrary statements stating that they should not be routinely offered, then both statements being discarded during round 3 – highlighting dissonance. During round 3 one final statement (‘respiratory exercises might be appropriate in some cases as part of rehabilitation’) was generated from open comments. Although, this final statement is more vague with regards to respiratory care, it may be appropriate in the context of other literature which suggests that the surgical approach chosen in addition to the stage of adolescence and growth mean that multiple factors will influence respiratory function (92, 95). Therefore, this dissonance warrants the need for further exploration of respiratory exercises as part of post-operative rehabilitation in AIS. Furthermore, aerobic and resistance training post-operatively have known benefits on respiratory function when compared to aerobic training alone (96). Clarity is needed on where these consensus statements regarding respiratory exercises fit with an aim of facilitating sports, exercise, and physical activity.

### Experts included within the study

Included within the surgical experts were three experts registered as physiatrists, medically qualified practitioners holding a medical license to practice. However, only one participant stated this at the start of the Delphi process. This study contained one open question asking participants to describe their practice, but did not explore the specific activities undertaken by experts and Delphi definitions of experts remains contentious and poorly defined (48). However, the literature discusses experts having the experience and knowledge of the issue and willingness and capacity to participate (48). Therefore, in discussion with the study steering group, it was determined that physiatrists, being medically qualified, met the definition of an expert, would be included in subsequent rounds and calculation of consensus. It was unclear from the data whether these participants had dual accreditation as both medical doctors and physiotherapists or whether their practice included a surgical component. However, it is widely accepted that many surgeons divide their time between activities such as clinics, teaching, and research (97). The proportion of surgeons who undertake operations and the portion of time spent doing so remains unknown (97). physiatrist responses were often more closely aligned with physiotherapist responses rather than surgeon responses, perhaps due to their training involving specific rehabilitation expertise (98). However, although physiatrist numbers are increasing, due to small numbers in this study it was not possible to demonstrate these differences with statistical testing (99).

## Strengths and Limitations

This international Delphi had several strengths, being the first study to establish expert surgeon and physiotherapy consensus on content of, philosophical approach, and MDT involvement in return to sports, exercise and physical activity following spinal fusion. Throughout this study all methodological considerations were discussed and agreed upon with patient and public involvement (PPI) representative (ER), and the study steering group (AS, NH, AG, AR). This study has been reported in accordance with Guidance on Conducting and REporting Delphi Studies guidelines as recommended by the Enhancing the QUAlity and Transparency Of Health Research network. All methods were described as per study protocol and subject to peer review process (33). Expert consensus has been established using inductive content analysis for the presence of themes, patterns or concepts followed by statistical testing to determine agreement and consensus in the group.

However, there were also some limitations to this Delphi study. All consenting eligible experts were able to participate regardless of whether they work part time or only work with conservatively managed individuals, this included 3 physiatrists within the surgical group although physiatrists as a unique profession was not individually sought. This study did not set out to involve other members of the MDT who may have valuable insight into post-operative rehabilitation. Due to small numbers of experts from the continents of Africa and South America and lacking information about hospital system, culture, and participant distribution it was not possible to evaluate dissonance globally. Finally, this study did not specify variables such as anterior or posterior surgical approach, or stage of growth when undergoing fusion which may influence components of rehabilitation, and return to sports, exercise, and physical activity.

### Implications for practice and research

This Delphi has successfully established an expert consensus from pre-operative care until 12 months post-operatively where there are no limitations on sports, exercise and physical activity participation. However, there remains a lack of detail, particularly regarding rehabilitation from 3 months post-operative onwards. Therefore, further development of consensus regarding the later phases of rehabilitation beyond 3 months post-operatively is required in the form of future pilot or feasibility testing of novel physiotherapeutic rehabilitation interventions for post-operative rehabilitation in AIS (36, 42, 50, 51). As discussed this Delphi highlighted areas of dissonance between experts, namely SSE and respiratory exercises, therefore future exploration of dissonance with experts may offer additional insights or reasons for diverging opinions such as demographic, cultural, or professional differences between individuals (34). A Delphi consensus does not replace original research and further development is required to form clinical guidelines regarding return to sports, exercise, and physical activity (51).

## Conclusion

This study was the first to generate an international consensus on return to sports, exercise, and physical activity following spinal fusion in AIS. A total of 71 consensus statements were generated that followed a series of themes that start with pre-operative care and end with final phase 4 rehabilitation and no restrictions at 12 months post-op. The latter stages of rehabilitation from intermediate to final phase lack detail with regards to returning to sports, exercise, and rehabilitation and warrant further development.

## Data Availability

All relevant data are within the manuscript and its Supporting Information files.

## Ethics and dissemination

Full ethical approval has been provided by the University of Birmingham, Reference number: ERN_1617-Nov2023. Dissemination will take place through conference presentation and peer reviewed publications.

## Author contributions

Study conceived by NH, AS, AG, AR, ST, with all authors contributing to shaping the design and methods. ST drafted the manuscript with guidance from AS. All authors contributed to the revising and re-drafting the manuscript. The final version was approved by all authors with agreement to submission to PloS One. AS is the guarantor of the work, with ST the corresponding author.

